# Awareness and Use of Sound- and Music-Based Gait Training in People with Parkinson’s Disease in Japan: A Cross-Sectional Survey

**DOI:** 10.64898/2025.12.15.25342289

**Authors:** Tomohiro Samma, Yuki Uno, Harukaze Yatsugi, Shinya Iseri, Yuki Matsuda, Toshiki Imagawa, Makoto Sawada, Shinya Fujii

## Abstract

Sound- and music-based gait training (SM-GT), including rhythmic auditory stimulation and music-based movement approaches, has been shown to improve gait and functional outcomes in people with Parkinson’s disease (PD). However, little is known about how such interventions are recognized and implemented in real-world settings. This study investigated awareness and actual use of SM-GT among people with PD in Japan across inpatient rehabilitation, home-based rehabilitation, and daily-life contexts. A cross-sectional questionnaire survey was conducted among 62 people with PD recruited from public lectures and exercise programs. Participants reported their awareness of SM-GT, experiences of use in different contexts, types of auditory cues employed, and functional status including activities of daily living, gait and balance, and freezing of gait. Overall, 57.4% of participants reported being aware of SM-GT, whereas actual use remained limited across contexts (26.1% in inpatient rehabilitation, 9.1% in home-based rehabilitation, and 31.1% in daily-life contexts). Awareness was higher among participants with inpatient rehabilitation experience than among those with home-based rehabilitation experience; however a substantial gap between awareness and actual use was observed across all settings. While metronomes were the most frequently recognized auditory cue, hand clapping or verbal cueing and music were more commonly used in practice. Awareness and use of SM-GT also varied according to functional status, with relatively higher implementation among individuals with mild functional impairment. These findings reveal a pronounced discrepancy between recognition and real-world implementation of SM-GT in Japan, highlighting the need for strategies that facilitate translation of evidence-based auditory interventions into routine rehabilitation and everyday walking for people with PD.

## Introduction

Parkinson’s disease (PD) is a progressive neurodegenerative disorder that impairs activities of daily living (ADL) and quality of life (QOL), and is characterized not only by motor symptoms such as resting tremor, rigidity, bradykinesia, and postural instability, but also by a wide range of non-motor symptoms, including sleep disturbances, depression, anxiety, cognitive decline, and autonomic dysfunction (Bock et al., 2022; Carapellotti et al., 2020; Gazewood et al., 2013; Li et al., 2024; Nemade et al., 2021; Peterson & Horak, 2016; Sveinbjornsdottir, 2016). With global population aging, the prevalence of PD has been increasing worldwide (GBD 2016 Parkinson’s Disease Collaborators, 2018; Pringsheim et al., 2014; Reeve et al., 2014). The World Health Organization reported that the number of people with PD exceeded 8.5 million in 2019—approximately double the estimates from 25 years earlier (World Health Organization, 2023). Recent projections further estimate that the global number of people with PD will reach approximately 25 million by 2050 (Mantri et al., 2025; Su et al., 2025), suggesting that PD is becoming an increasingly critical public health concern (Ben-Shlomo et al., 2024; Dorsey & Bloem, 2018; Su et al., 2025).

Among the diverse symptoms of PD, gait disturbances are of particular clinical importance. Gait abnormalities such reduced stride length, decreased walking speed, and increased gait variability have been shown to be strongly associated with an elevated fall risk, reduced mobility, and declines in both ADL and QOL (Bradley et al., 2025; Creaby & Cole, 2018; Smulders et al., 2016). Freezing of gait (FoG) is a unique and disabling clinical phenomenon characterized by brief episodic absence or marked reduction of forward progression of the feet despite the intention to walk. These episodes typically occur during gait initiation or turning and may be accompanied by extremely short steps or a sensation of “glued ” feet (Nutt et al., 2011). FoG affects approximately 38.2–81% of people with PD as the disease progresses (Hely et al., 2008; Perez-Lloret et al., 2014). FoG constitutes a severe motor symptom closely linked to falls, avoidance of outdoor activities, and reductions in health-related QOL (Cronin et al., 2024; Ge et al., 2020; Gilat et al., 2021; Herman et al., 2023; Nutt et al., 2011; Perez-Lloret et al., 2014).

Although pharmacological treatments such as levodopa and deep brain stimulation are effective for many motor symptoms of PD, gait disturbances and FoG often show insufficient or inconsistent therapeutic responses, and are therefore regarded as major treatment-resistant symptoms of PD (Delgado-Alvarado et al., 2020; Gilat et al., 2021; Herman et al., 2023).

In light of these challenges, the importance of non-pharmacological rehabilitation strategies—such as gait training and exercise therapy using visual, auditory, or somatosensory cues—has been repeatedly emphasized. Among these, movement interventions employing auditory cues, including rhythmic auditory stimulation (RAS) and music-based approaches, have been shown to improve gait parameters such as walking speed and stride length (Ashoori et al., 2015; Nieuwboer et al., 2007; Nombela et al., 2013; Thaut et al., 1996), as well as to exert beneficial effects on functional outcomes including balance, mobility, ADL, and QOL (Forte et al., 2021; Wang et al., 2022; Ye et al., 2022). Subsequent studies and meta-analyses have similarly reported improvements in gait and functional indicators, such as the six-minute walk distance and performance on the Timed Up and Go test (Bella et al., 2017; Ghai et al., 2018; Wang et al., 2022; Ye et al., 2022). Interventions involving clapping or verbal cueing by therapists or caregivers have also been suggested to act as external cues embedded within interpersonal interaction, potentially enhancing attentional focus and motivation (Ashoori et al., 2015; Rocha et al., 2014). More recently, RAS delivered through wearable devices or digital applications has been reported to support improvements in daily-life walking and home-based rehabilitation (Bourdon et al., 2025; Cochen De Cock et al., 2021; Ginis et al., 2017; Porciuncula et al., 2025; Scataglini et al., 2025). Music-based movement therapy (MbM), which involves the use of music itself and encompasses approaches such as gait and balance training or dance based on musical beat structures, represents a broader concept and has demonstrated significant effects on gait ability, balance (Bella et al., 2015; Calabrò et al., 2019; Thaut et al., 2019), and QOL (Pohl et al., 2020). Beyond compensating for the temporal organization of movement via rhythmic cues, MbM uniquely incorporates elements less prominent in conventional exercise therapies, such as enjoyment, enhanced motivation, and emotional regulation associated with music listening (Bella et al., 2017; Lee & Ko, 2023; Nombela et al., 2013).

Taken together, these findings suggest that external auditory cues—particularly interventions using sound or music—represent a promising approach for gait rehabilitation among people with PD. However, much of the existing research has focused on evaluating the efficacy of such interventions through clinical trials or laboratory-based studies, and far less is known about the extent to which “sound- and music-based gait training (SM-GT)” is actually implemented in clinical practice or daily life, as well as how it is perceived by people with PD. Outside the context of gait rehabilitation, previous work has examined the use of music in everyday life among people with PD and their responsiveness to musical rhythm (Nombela et al., 2013), and more recent online surveys have investigated the purposes for which people with PD use music in daily life, as well as its emotional and motivational effects (Rose et al., 2023, 2025).

Nevertheless, quantitative research that focuses specifically on awareness of SM-GT and on its use in rehabilitation settings and everyday walking remains limited.

Against this background, the present study aimed to examine awareness and actual use of SM-GT among people with PD in Japan. In this study, SM-GT broadly refers to interventions in which rhythmic auditory cues—such as metronome beats, hand clapping, verbal cues, or music—are used during walking to support the regulation and stabilization of gait rhythm and to facilitate the execution of walking. Specifically, the study sought to describe the extent to which SM-GT is recognized, as well as how it is used across different contexts, including inpatient rehabilitation, home-based rehabilitation, and daily life. In addition, we investigated the types of auditory cues currently employed, thereby distinguishing between those that are recognized as part of SM-GT and those that are actually used in practice. Furthermore, by exploring associations with functional status—including ADL level, gait and balance status, and the severity of FoG—we aimed to clarify how awareness and use of SM-GT are distributed across different functional profiles of people with PD. By elucidating real-world patterns of awareness and use, this study offers a complementary perspective to existing clinical trials that have demonstrated the efficacy of SM-GT, and contributes to considerations of how SM-GT might be more effectively implemented and disseminated in clinical and everyday contexts.

## Methods

### Participants

Approval for this study was obtained from the Research Ethics Committee of Reiwa Health Sciences University, and the study was conducted in accordance with the principles of the Declaration of Helsinki. Written informed consent was obtained from all participants before participation.

Participants were recruited during public lectures and exercise classes organized for people with PD at three locations in Japan: Kobe, Akashi, and Yonago. All participants had received a clinical diagnosis of PD from a physician. The questionnaire was distributed to and collected from participants immediately after the lectures and exercise classes concluded.

### Questionnaires

The survey was administered in a paper-based format following public lectures and exercise classes conducted for people with PD. Before participants began the questionnaire, a trained researcher delivered standardized instructions to all respondents simultaneously.

### Background information

Participants first reported demographic and clinical information, including date of birth, sex, prefecture of residence, onset date of PD, date of clinical diagnosis, comorbid medical conditions, and current medication status.

Activities of daily living (ADL) status was assessed using the Modified Rankin Scale (mRS; (van Swieten et al., 1988)), administered as a self-report questionnaire based on the Japanese version of the scale (Matsuda et al., 2025). Participants were asked to select the category that best described their usual daily functioning in the absence of medication effects. Responses were classified into mRS scores from 0 to 4 according to the original definitions: score 0 (*asymptomatic*; no symptoms at all), score 1 (*symptomatic without disability*; able to carry out all usual activities), score 2 (*mild disability*; unable to perform all previous activities but able to manage personal affairs without assistance), score 3 (*moderate disability*; requiring some help but able to walk without assistance), and score 4 (*moderately severe disability*; unable to walk or attend to bodily needs without assistance). Severity descriptors were used solely for descriptive clarity; classification strictly followed the original mRS definitions. The categories of severe disability (score 5) and death (score 6) were not included in the present study.

Gait and balance status were assessed using Item 2.12 (walking and balance) of the MDS-UPDRS (Goetz et al., 2008). Participants were classified into five categories according to the original item definitions. Specifically, the *Normal* category indicated no problems with walking or balance. The *Slight* category included individuals who were slightly slow or dragged a leg but never used a walking aid. The *Mild* category referred to occasional use of a walking aid without requiring assistance from another person. The *Moderate* category indicated usual use of a walking aid (e.g. cane or walker) while remaining independent of personal assistance. The *Severe* category denoted the need for support from another person to walk safely.

FoG was evaluated using the Item 2.13 (Freezing) of the MDS-UPDRS (Goetz et al., 2008). Participants were classified according to the original item definitions. The *Normal* category indicated no freezing episodes. The *Slight* category referred to brief freezing episodes from which individuals could easily resume walking, without requiring assistance from another person or the use of a walking aid. The *Mild* category denoted freezing accompanied by difficulty in gait initiation, although assistance from another person or a walking aid was not required. The *Moderate* category indicated freezing that caused substantial difficulty in restarting gait and sometimes necessitated the use of a walking aid or assistance from another person. The *Severe* category described freezing that required the use of a walking aid or assistance from another person most or all of the time.

### Items regarding sound- and music-based gait training (SM-GT)

After completing the background section, participants responded to items concerning SM-GT. In this study, SM-GT was defined as a method in which rhythmic auditory cues (e.g., metronome beats or hand claps) are presented during walking to improve or stabilize gait rhythm, or as a method in which individuals walk in synchrony with the metrical structure of music to support gait tempo regulation and walking execution.

Participants were first asked about their awareness of SM-GT. Those who indicated awareness were further asked about the types of auditory cues they recognized and the contexts in which these auditory cues had been used. Participants were then surveyed separately about their experiences with SM-GT in inpatient rehabilitation and in home-based rehabilitation provided by physical or occupational therapists. These sections asked about the frequency of gait training without auditory cues, whether participants had experience with SM-GT, the types and frequency of auditory cues used when applicable, and perceived differences in effectiveness compared with gait training without auditory cues. Finally, participants were asked about their use of auditory cues during walking in daily life. They indicated whether they used auditory cues to support walking. Those who responded affirmatively reported the types of auditory cues used, frequency of use, whether they continued using such cues, perceived effectiveness in everyday contexts, and the sources of the auditory cues.

Details of all questionnaire items are provided in supporting information.

### Data analysis

For awareness and use of SM-GT, the number and proportion of responses were calculated, and chi-square goodness-of-fit tests were conducted to examine whether the response distributions differed significantly from an equal distribution (50% vs. 50%). Effect sizes were quantified using Cohen’s w. To examine associations with participants’ functional status, the number and proportion of participants reporting awareness or use of SM-GT were calculated separately for each category of ADL level, gait status, and FoG, and the results were summarized in contingency tables. Because some functional categories included small sample sizes, comparisons involving these groups were reported primarily using descriptive statistics.

In addition, among participants who reported being aware of SM-GT or having experience using it, responses regarding the types of auditory cues recognized or used (metronome, music, hand clapping/verbal cueing) were aggregated, and the number and proportion of responses for each category were calculated. As multiple responses were permitted for the auditory cue items, the total number of responses could exceed the number of respondents.

For all analyses, data from participants who did not respond to the relevant items were excluded from the corresponding analyses. The significance level was set at α = 0.05 (two-tailed) for all statistical tests. All statistical analyses were conducted using R. The dataset collected in this study has been made publicly available in the Open Science Framework (OSF) repository at https://osf.io/5v2fu/.

## Results

### Sample Characteristics

Initially, written informed consent and completed questionnaires were obtained from 64 individuals (39 women, 24 men, and 1 with unspecified sex). Two respondents were excluded due to missing data on key items, resulting in a final analytic sample of 62 participants (37 women, 24 men, and 1 with unspecified sex). The mean age of the analytic sample was 71.34 years (SD = 7.83; range = 47–90 years).

Regarding ADL, 46.8% of participants were classified as *Mild disability* (n = 29), 24.2% as *Symptomatic without disability* (n = 15), 22.6% as *Moderate disability* (n = 14), and 3.2% each as *Asymptomatic* and *Moderately severe disability* (n = 2 for both). Regarding gait and balance status, 45.2% of participants were classified as *Slight* (n = 28), 30.7% as *Moderate* (n = 19), 12.9% as *Normal* (n = 8), 8.1% as *Mild* (n = 5), and 3.2% as *Severe* (n = 2). Regarding FoG status, 35.5% of participants were classified as *Normal* (n = 22), 32.3% as *Slight* (n = 20), 17.7% as *Moderate* (n = 11), 12.9% as *Mild* (n = 8), and 1.6% as *Severe* (n = 1).

Among the respondents, 23 individuals (37.1%; 13 women, 9 men, and 1 with unspecified sex) reported having experience with inpatient rehabilitation (Figure 1). Regarding ADL status, 43.5% were classified as *Mild disability* (n = 10), 39.1% as *Moderate disability* (n = 9), 13.0% as *Symptomatic without disability* (n = 3), and 4.3% as *Moderately severe disability* (n = 1). For gait and balance status, the most common categories were *Slight* (34.8%, n = 8) and *Moderate* (34.8%, n = 8), followed by *Mild* (13.0%, n = 3), *Severe* (8.7%, n = 2), and *Normal* (8.7%, n = 2). Regarding FoG status, the most frequent category was *Moderate* (34.8%, n = 8), followed by *Normal* (26.1%, n = 6), *Mild* (21.7%, n = 5), *Slight* (13.0%, n = 3), and *Severe* (4.3%, n = 1).

**Figure 1.**
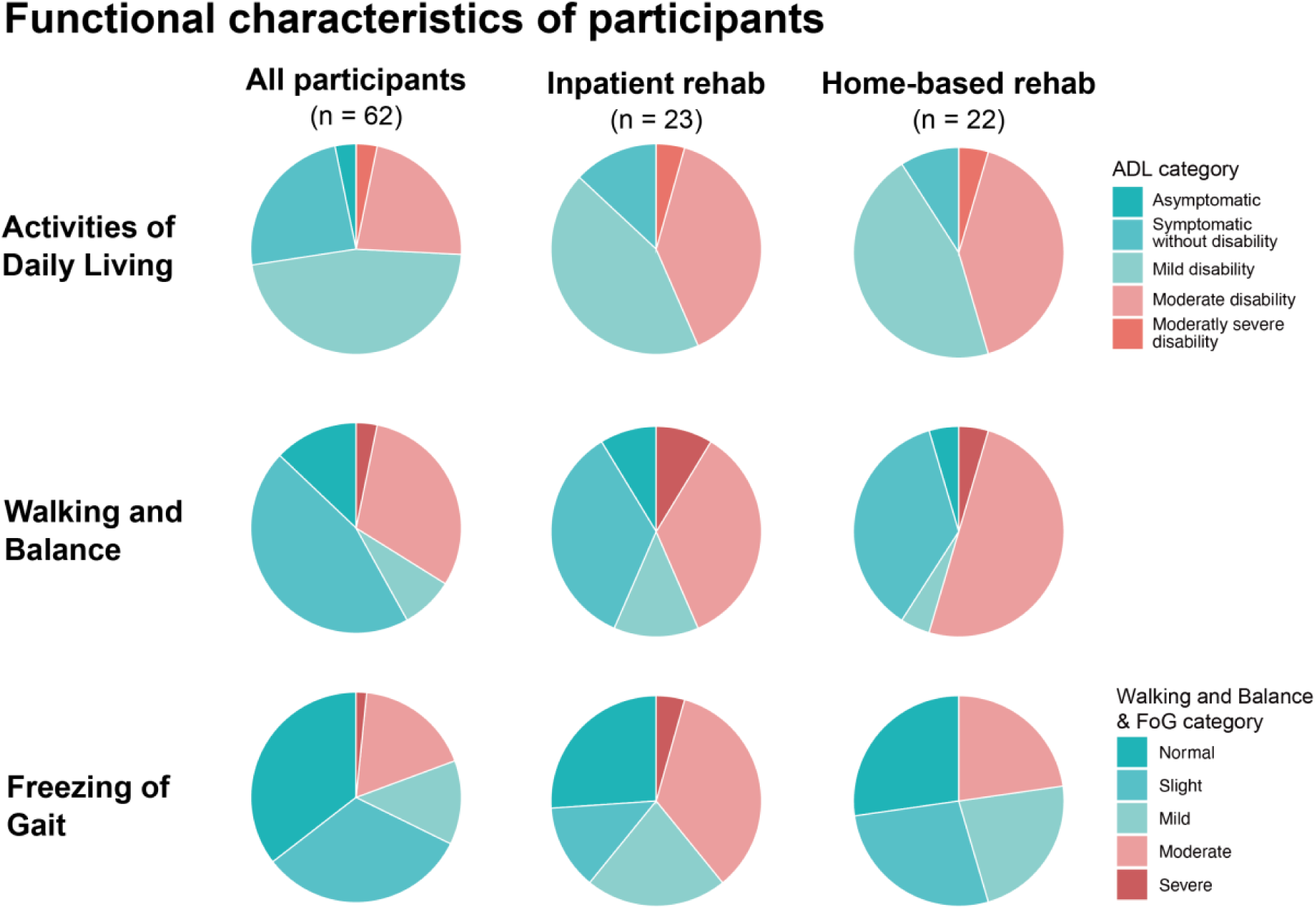
Functional characteristics of participants by rehabilitation experience. Pie charts show the distribution of functional status across three domains: activities of daily living (ADL), gait and balance status, and freezing of gait (FoG) severity. Distributions are presented for the overall sample (left column), participants with inpatient rehabilitation experience (middle column), and participants with home-based rehabilitation experience (right column). Colors represent increasing functional severity from normal (blue) to severe (red).

Among the respondents, 22 individuals (35.5%; 14 women and 8 men) reported having experience with gait training during home-based rehabilitation conducted by professionals such as physical or occupational therapists (Figure 1). Regarding ADL status, the most common classifications were *Mild disability* (45.5%, n = 10), followed by *Moderate disability* (41.0%, n = 9), *Symptomatic without disability* (9.0%, n = 2) and *Moderately severe disability* (4.5%, n = 1). For gait and balance status, the most frequent classification was *Moderate* (50.0%, n = 11), followed by *Slight* (36.4%, n = 8). *Mild*, *Severe*, and *Normal* were each reported by 4.5% of participants (n = 1 for each category). Regarding FoG status, *Normal* and *Slight* were the most common category (both 27.3%, n = 6), followed by *Moderate*, and *Mild*, each at 22.7% (n = 5), and no participants were classified as *Severe*.

### Awareness of SM-GT

Of the respondents, 57.4% (n = 35) reported being aware of SM-GT, whereas 42.6% (n = 26) reported being unaware of it (valid responses = 61; one nonresponse; Figure 2 and 6). A chi-square goodness-of-fit test was conducted to determine whether the response distribution deviated from an equal 50%–50% split, and no significant deviation was observed (*χ²*(1) = 1.33, *p* = 0.25, *Cohen’s w* = 0.15). Among participants who were aware of SM-GT (n = 35), a multiple-response question about the types of auditory cues they recognized showed that metronome sounds were selected most frequently (n = 27), followed by music (n = 21) and hand clapping or verbal cues (n = 18).

**Figure 2.**
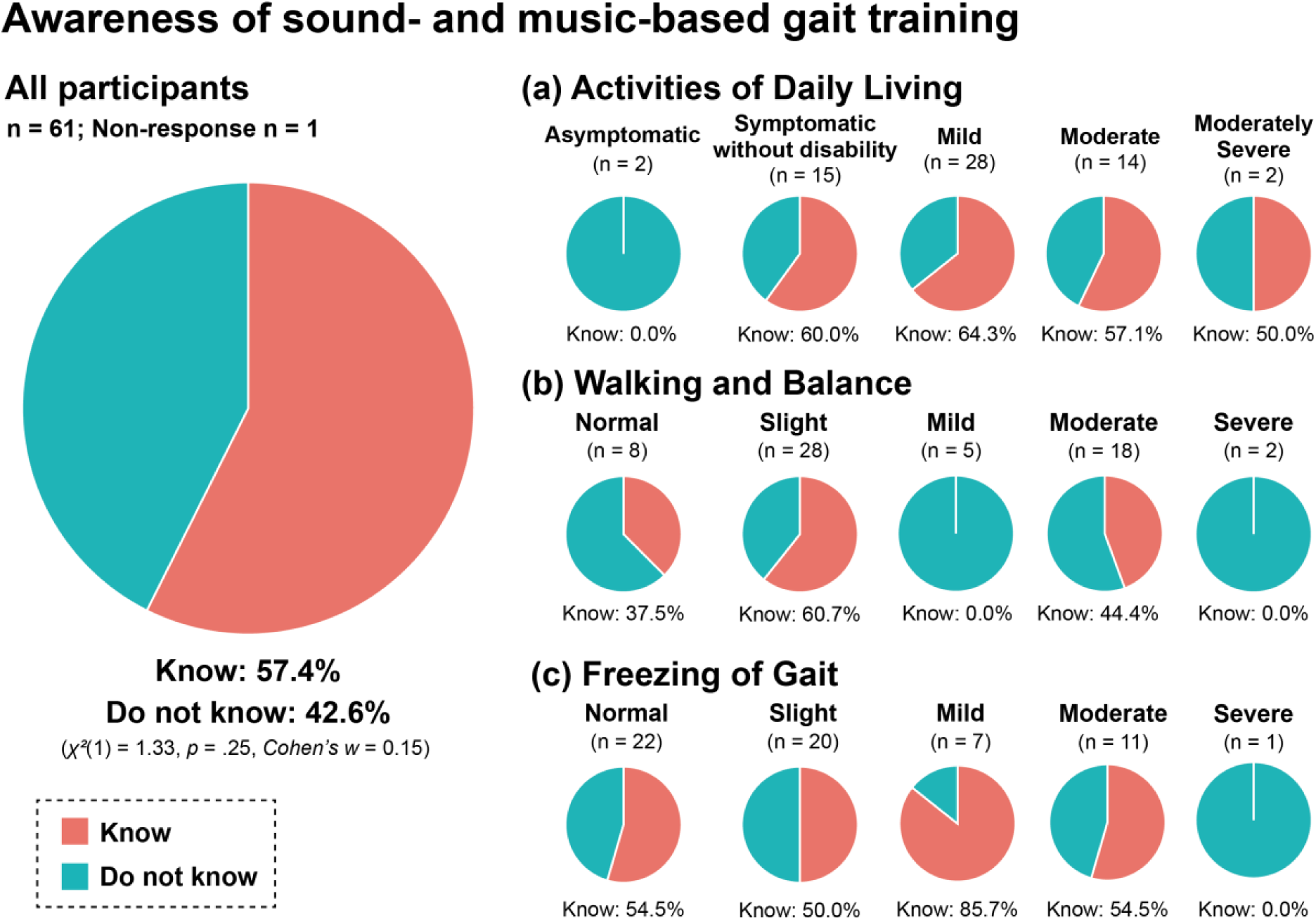
Awareness of sound- and music-based gait training (SM-GT) among participants. Pie charts illustrate the proportion of participants who reported being aware of SM-GT (“Know”) versus not aware (“Do not know”). The large chart on the left shows overall awareness among all respondents (n = 61; non-response = 1). The smaller charts display awareness stratified by Activities of Daily Living (ADL) level, Walking and Balance status, and Freezing of Gait (FoG) severity. Percentages represent the proportion of participants within each subgroup who reported being aware of SM-GT.

Based on participants’ ADL levels, awareness of SM-GT was compared (Figure 2a). In the *Asymptomatic* category (n = 2), 100% (n = 2) reported being unaware, indicating that no participants were aware of SM-GT. In the *Symptomatic without disability* category (n = 15), 60.0% (n = 9) reported awareness, reflecting a moderate level of recognition. In the *Mild disability* category (n = 28; 1 nonresponse), 64.3% (n = 18) reported awareness, representing the highest proportion among the five ADL levels. In the *Moderate disability* category (n = 14), 57.1% (n = 8) reported awareness, a proportion not markedly different from that observed in the *Mild disability* category. In the *Moderately severe disability* category (n = 2), 50.0% (n = 1) reported awareness.

Awareness of SM-GT was also examined across gait and balance status (Figure 2b). In the *Normal* category (n = 8), 62.5% (n = 5) were unaware and 37.5% (n = 3) were aware. In the *Slight* category (n = 28), 39.3% (n = 11) were unaware and 60.7% (n = 17) were aware, indicating a relatively higher level of recognition. In contrast, all participants in the *Mild* category (n = 5) reported being unaware. In the *Moderate* category (n = 18; 1 nonresponse), 55.6% (n = 10) were unaware and 44.4% (n = 8) were aware, showing a relatively balanced distribution. In the *Severe* category (n = 2), 100% (n = 2) reported being unaware.

Awareness of SM-GT was further compared based on FoG severity (Figure 2c). In the *Normal* category (n = 22), 54.5% (n = 12) reported awareness and 45.5% (n = 10) reported unawareness, indicating an approximately even distribution. A similar pattern was observed in the *Slight* category (n = 20), in which awareness and unawareness were evenly split at 50.0% (n = 10) each. In the *Mild* category (n = 7; 1 nonresponse), 85.7% (n = 6) reported awareness. In the *Moderate* category (n = 11), 54.5% (n = 6) were aware and 45.5% (n = 5) were unaware, a distribution comparable to that of the *Normal* category. In contrast, the sole participant classified as *Severe* (n = 1) reported being unaware of SM-GT.

### Inpatient Rehabilitation: Awareness, Use, and Frequency of SM-GT

Among the 23 participants who had experience with inpatient rehabilitation, 52.2% (n = 12) reported engaging in gait rehabilitation “daily.” Additionally, 8.7% (n = 2) reported engaging in gait rehabilitation “two to three times per week,” 8.7% (n = 2) reported “once per week,” and 4.3% (n = 1) reported “once every two weeks.” A further 26.1% (n = 6) indicated participating at a lower frequency.

Among participants with inpatient rehabilitation experience (n = 23), 73.9% (n = 17) reported being aware of SM-GT, whereas 26.1% (n = 6) reported being unaware (Figure 6). A chi-square goodness-of-fit test comparing the response distribution with an equal 50%–50% split indicated a significant deviation, showing that the proportion of participants who were aware of SM-GT was significantly higher than 50% (*χ²*(1) = 5.3, *p* = 0.02, *Cohen’s w* = 0.48). Of those with inpatient rehabilitation experience, 26.1% (n = 6) reported having used SM-GT (Figure 3). A chi-square goodness-of-fit test on the distribution of SM-GT use also revealed a significant deviation from equality, with the proportion of users being significantly lower than 50% (*χ²*(1) = 5.3, *p* = 0.02, *Cohen’s w* = 0.48). A multiple-response question regarding the types of auditory cues used showed that four participants had used metronome sounds, four had used hand clapping or verbal cues, and three had walked in synchrony with music. Among the six participants who engaged in SM-GT during hospitalization, 16.7% (n = 1) reported doing so “daily,” 33.3% (n = 2) “two to three times per week,” 16.7% (n = 1) “once per week,” and 33.3% (n = 2) “less than once every two weeks.” Participants were also asked how effective they perceived SM-GT to be compared with gait training without auditory cues (n = 5; one nonresponse). Of these, 40.0% (n = 2) reported that they “felt the effects,” another 40.0% (n = 2) reported “feeling the effects to some extent,” while 20.0% (n = 1) reported “not feeling much effect.”

**Figure 3.**
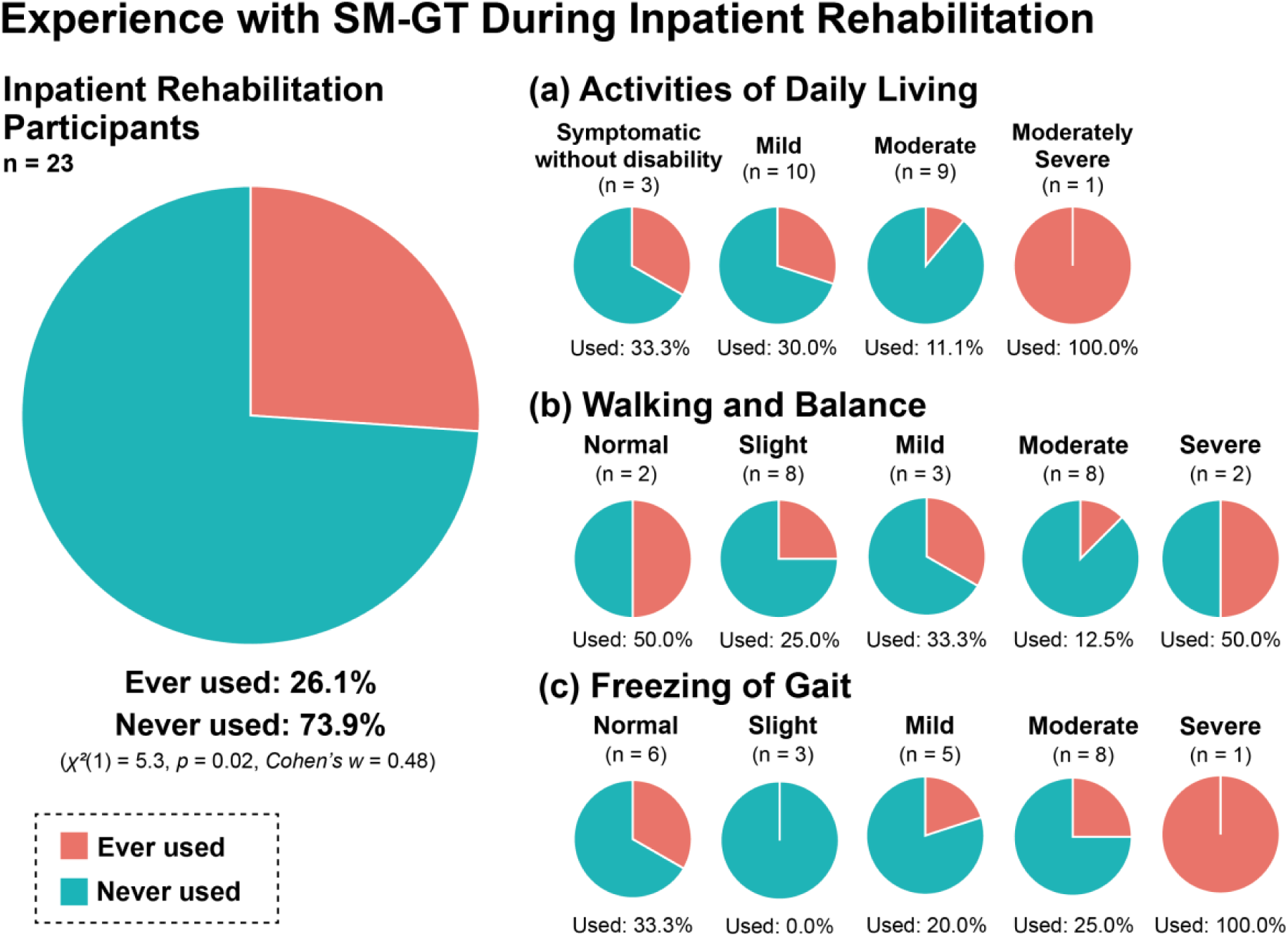
Experience with sound- and music-based gait training (SM-GT) during inpatient rehabilitation. Pie charts illustrate the proportion of participants who reported having used SM-GT (“Ever used”) versus never used (“Never used”) during inpatient rehabilitation. The large chart on the left shows overall SM-GT use among participants with inpatient rehabilitation experience (n = 23). The smaller charts display usage stratified by Activities of Daily Living (ADL) level, Walking and Balance status, and Freezing of Gait (FoG) severity. Percentages represent the proportion of participants within each subgroup who reported having used SM-GT.

Experience with SM-GT during inpatient rehabilitation was examined based on participants’ ADL levels (Figure 3a). In the *Symptomatic without disability* category (n = 3), 33.3% (n = 1) reported having experience with SM-GT during hospitalization. In the *Mild disability* category (n = 10), 30.0% (n = 3) reported such experience. In the *Moderate disability* category (n = 9), 11.1% (n = 1) reported having experience with SM-GT, whereas in the *Moderately severe disability* category (n = 1), 100.0% (n = 1) reported experience. Experience with SM-GT during inpatient rehabilitation was also examined based on gait and balance status (Figure 3b). In the *Normal* category (n = 2), 50.0% (n = 1) reported experience, while in the *Slight* category (n = 8), 25.0% (n = 2) reported experience. In the *Mild* category (n = 3), 33.3% (n = 1) reported experience, and in the *Moderate* category (n = 8), 12.5% (n = 1) reported experience. In the *Severe* category (n = 2), 50.0% (n = 1) reported experience with SM-GT during hospitalization. Finally, experience with SM-GT during inpatient rehabilitation was examined according to FoG severity (Figure 3c). In the *Normal* category (n = 6), 33.3% (n = 2) reported experience, while in the *Slight* category (n = 3), no participants reported experience (0.0%). In the *Mild* category (n = 5), 20.0% (n = 1) reported experience, and in the *Moderate* category (n = 8), 25.0% (n = 2) reported experience. In contrast, in the *Severe* category (n = 1), 100.0% (n = 1) reported having experience with SM-GT during inpatient rehabilitation.

### Home-based Rehabilitation: Awareness, Use, and Frequency of SM-GT

Among the 22 participants who had experience with home-based rehabilitation provided by physical or occupational therapists, 4.5% (n = 1) reported performing gait rehabilitation “daily.” Additionally, 18.2% (n = 4) reported performing it “two to three times per week,” 40.9% (n = 9) “once per week,” and 31.8% (n = 7) at a lower frequency. One participant (4.5%) did not provide a response.

Among participants with home-based rehabilitation experience (n = 21; one nonresponse), 47.6% (n = 10) reported being aware of SM-GT, whereas 52.4% (n = 11) reported being unaware (Figure 6). A chi-square goodness-of-fit test comparing the response distribution with an equal 50%–50% split indicated no significant deviation (χ²(1) = 0.05, p = 0.83, Cohen’s w = 0.05). Among those who had home-based rehabilitation experience (n = 22), 9.1% (n = 2) reported having used SM-GT (Figure 4). A chi-square goodness-of-fit test indicated a significant deviation from equality (χ²(1) = 16.7, p < 0.01, Cohen’s w = 0.83), showing that the proportion of SM-GT users was significantly lower than 50%. A multiple-response question regarding the types of auditory cues used revealed that one participant had used metronome sounds, two had used hand clapping or verbal cues, and one had walked in synchrony with music. Among the two participants who practiced SM-GT at home, 50.0% (n = 1) reported doing so “once every two weeks,” and 50.0% (n = 1) reported practicing it “less than once every two weeks.” When asked about the perceived effectiveness of SM-GT compared with gait training without auditory cues, both participants (100.0%, n = 2) reported “feeling the effects to some extent.”

**Figure 4.**
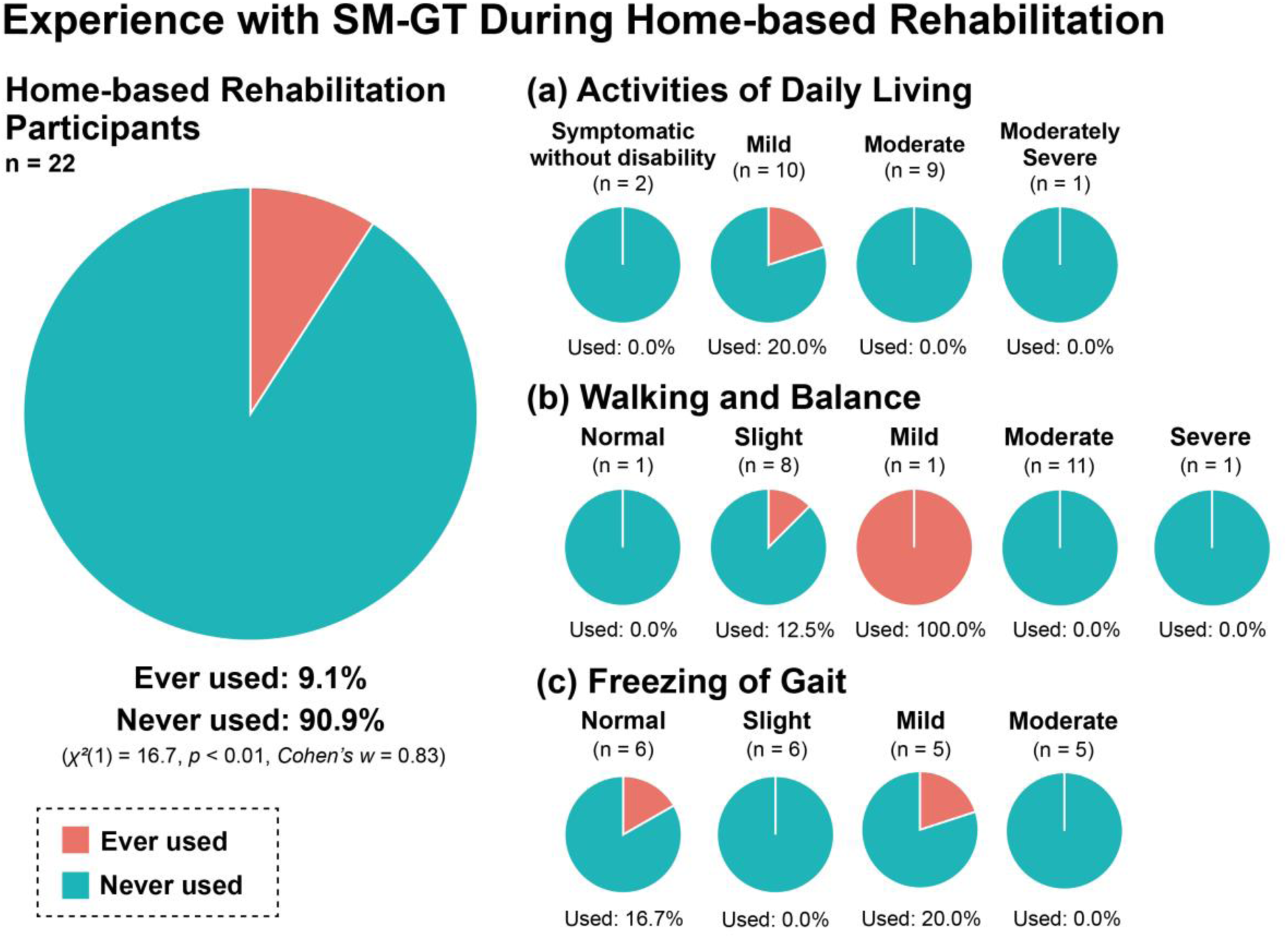
Experience with sound- and music-based gait training (SM-GT) during home-based rehabilitation. Pie charts illustrate the proportion of participants who reported having used SM-GT (“Ever used”) versus never used (“Never used”) during home-based rehabilitation. The large chart on the left shows overall SM-GT use among participants with home-based rehabilitation experience (n = 22). The smaller charts display usage stratified by Activities of Daily Living (ADL) level, Walking and Balance status, and Freezing of Gait (FoG) severity. Percentages represent the proportion of participants within each subgroup who reported having used SM-GT.

Experience with SM-GT during home-based rehabilitation was examined based on ADL levels (Figure 4a). Only participants in the *Mild disability* category (n = 10) reported experience, with 20.0% (n = 2) indicating that they had practiced SM-GT at home. No participants in the *Symptomatic without disability* category (n = 2), *Moderate disability* category (n = 9), or *Moderately severe disability* category (n = 1) reported experience. Experience with SM-GT during home-based rehabilitation was also examined according to gait and balance status (Figure 4b). In the Slight category (n = 8), 12.5% (n = 1) reported experience, and in the Mild category (n = 1), 100.0% (n = 1) reported experience. However, no participants reported experience in the Normal category (n = 1), Moderate category (n = 11), or Severe category (n = 1). Finally, experience with SM-GT during home-based rehabilitation was examined based on FoG severity (Figure 4c). In the Normal category (n = 6), 16.7% (n = 1) reported experience, and in the Mild category (n = 5), 20.0% (n = 1) reported experience. In contrast, no participants reported experience in the Slight category (n = 6) or Moderate category (n = 5).

### Daily-Life Use of SM-GT

Among the 61 valid responses (one nonresponse), 31.1% (n = 19) of participants reported having used SM-GT in daily life, whereas 68.9% (n = 42) reported no such experience (Figure 5 and 6). A chi-square goodness-of-fit test comparing this distribution with an equal 50%–50% split indicated a significant deviation (*χ²*(1) = 8.7, *p* < 0.01, *Cohen’s w* = 0.38), showing that the rate of daily-life use was significantly lower than 50%.

**Figure 5.**
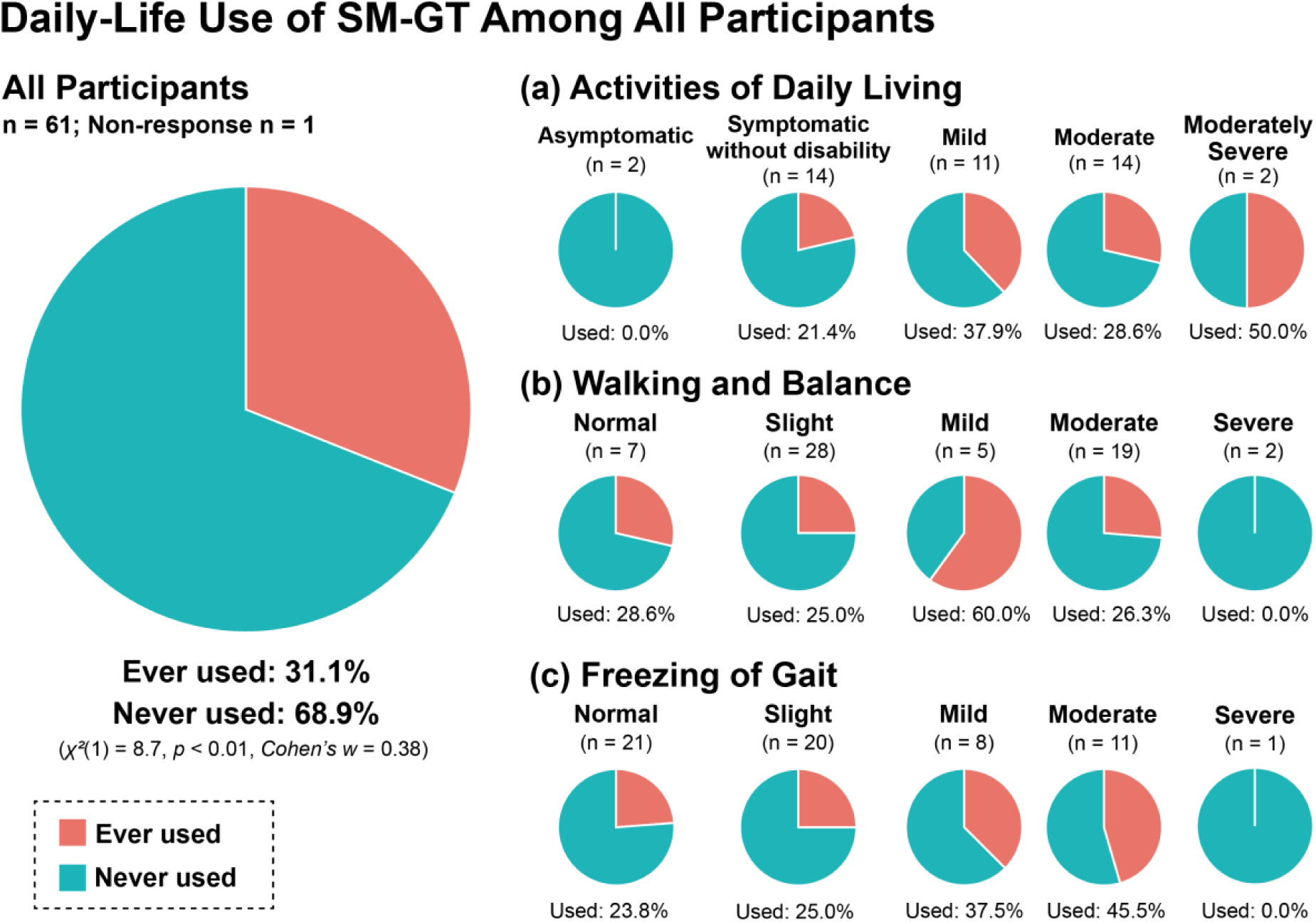
Daily-life use of sound- and music-based gait training (SM-GT) among participants. Pie charts illustrate the proportion of participants who reported using SM-GT (“Ever used”) versus never using it (“Never used”) in daily life. The large chart on the left shows overall SM-GT use among all respondents (n = 61; 22 reported use). The smaller charts display usage stratified by Activities of Daily Living (ADL) level, Walking and Balance status, and Freezing of Gait (FoG) severity. Percentages represent the proportion of participants within each subgroup who reported using SM-GT in daily life.

**Figure 6.**
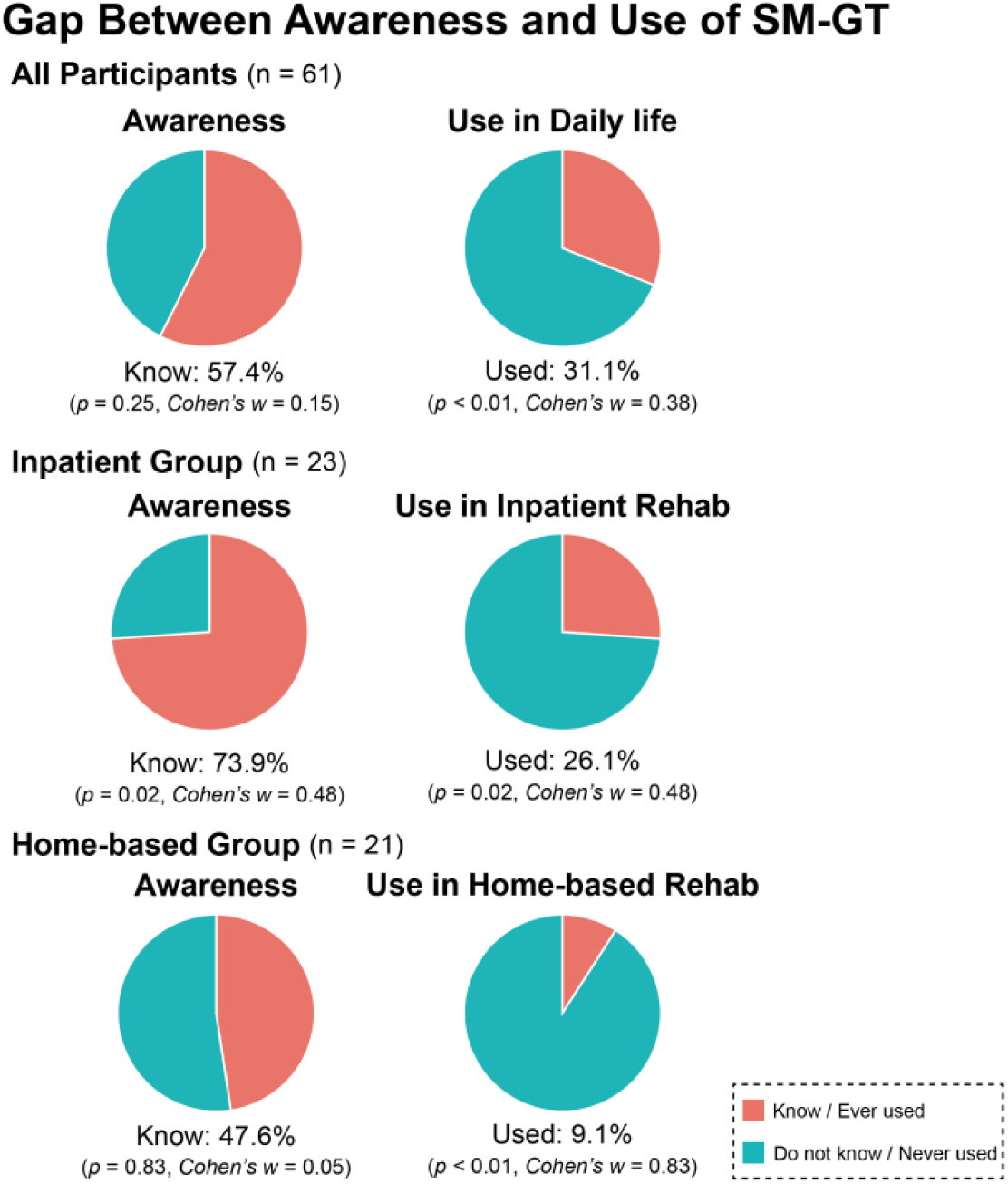
Gap between awareness and use of sound- and music-based gait training (SM-GT). Pie charts illustrate the proportions of participants who reported being aware of SM-GT (“Know”) and those who had experience using SM-GT (“Ever used”) across three contexts: the overall sample (n = 61), participants with inpatient rehabilitation experience (n = 23), and those with home-based rehabilitation experience (n = 21). Within each group, awareness (left) and use (right) are displayed side by side to highlight the discrepancy between recognition and actual implementation. Awareness of SM-GT was moderate to high across groups (57.4–73.9%), whereas usage remained substantially lower in all contexts (9.1–31.1%). Reported p-values reflect chi-square goodness-of-fit tests comparing observed distributions with an equal 50:50 expectation, and Cohen’s w provides effect-size estimates for deviation from this distribution.

Daily-life use of SM-GT was then examined based on participants’ ADL levels (Figure 5a). In the *Asymptomatic* category (n = 2), no participants reported daily-life use. In the *Symptomatic without disability* category (n = 14; one nonresponse), 21.4% (n = 3) reported using SM-GT in daily life. In the *Mild disability* category (n = 29), 37.9% (n = 11) reported daily-life use, representing the highest proportion among the five ADL categories. In the *Moderate disability* category (n = 14), 28.6% (n = 4) reported use. In the *Moderately severe disability* category (n = 2), 50.0% (n = 1) reported daily-life use.

Daily-life use of SM-GT was also examined according to gait and balance status (Figure 5b). In the *Normal* category (n = 7; one nonresponse), 28.6% (n = 2) reported use. In the *Slight* category (n = 28), 25.0% (n = 7) reported use, with the majority reporting no experience. In the *Mild* category (n = 5), 60.0% (n = 3) reported daily-life use, representing a relatively high proportion. In the *Moderate* category (n = 19), 26.3% (n = 5) reported use. In the *Severe* category (n = 2), no participants reported daily-life use.

Daily-life use of SM-GT was further examined based on FoG severity (Figure 5c). In the *Normal* category (n = 21; one nonresponse), 23.8% (n = 5) reported use. In the *Slight* category (n = 20), 25.0% (n = 5) reported use, a proportion similar to that of the *Normal* category. In the *Mild* category (n = 8), 37.5% (n = 3) reported daily-life use. In the *Moderate* category (n = 11), 45.5% (n = 5) reported use, representing the highest proportion among the FOG categories. In contrast, the sole participant in the *Severe* category (n = 1) reported no daily-life use of SM-GT. Among participants who reported using SM-GT in daily life, a multiple-response question revealed that hand clapping or verbal cues were most frequently used (n = 10), followed by music (n = 8) and metronome sounds (n = 2).

## Discussion

The present study aimed to clarify awareness of SM-GT and its actual use across inpatient rehabilitation, home-based rehabilitation, and daily-life contexts among people with PD in Japan. We further sought to examine the types of auditory cues employed and to explore associations between awareness and use of SM-GT and functional characteristics, including ADL level, gait and balance status, and FoG severity. The results showed that 57.4% of all participants reported being aware of SM-GT, with higher awareness among those with inpatient rehabilitation experience (73.9%) than among those with home-based rehabilitation experience (47.6%). In contrast, actual use of SM-GT was limited across contexts, with usage rates of 26.1% in inpatient rehabilitation, 9.1% in home-based rehabilitation, and 31.1% in daily life. These findings indicate a substantial gap between awareness and use of SM-GT (Figure 6). Exploratory analyses further revealed that awareness and use of SM-GT were unevenly distributed according to functional status, with certain biases observed across levels of ADL, gait and balance status, and FoG severity. In addition, although the metronome was the most frequently recognized auditory cue associated with SM-GT, hand clapping and verbal cueing were the most commonly used cues in practice. This discrepancy highlights a further gap between the auditory cues that are recognized as part of SM-GT and those that are actually implemented.

### Overall Awareness of SM-GT and Differences by Functional Status

In the present study, overall awareness of SM-GT among people with PD in Japan was 57.4%, indicating that approximately half of the participants were aware of its existence. Although previous studies have reported people with PD’s use of music and their perceived symptom-related benefits of music (Nombela et al., 2013; Rose et al., 2023, 2025), very few studies have specifically addressed awareness of SM-GT itself. In this regard, the present findings provide foundational evidence that helps fill this gap.

A further notable finding was that awareness of SM-GT varied according to functional status, including ADL level, gait status, and FoG severity. When examined by ADL level, awareness of SM-GT was approximately 50–65% across all groups except the *Asymptomatic* group. This pattern is consistent with previous reports indicating that many non-pharmacological rehabilitation and exercise programs primarily target individuals with mild to moderate PD who retain independent walking ability (Barnish & Barran, 2020; Nieuwboer et al., 2007; Schenkman et al., 2012). Moreover, it has been suggested that patients with *Moderate* to *Moderately severe* disability often experience insufficient access to necessary rehabilitation services and allied health professionals (Nielsen et al., 2022; Zaman et al., 2021). The relatively lower awareness observed in the more severe groups in the present study may therefore reflect such disparities in access to healthcare and rehabilitation services.

Comparisons based on gait status revealed that awareness of SM-GT was highest in the *Slight* group, characterized by mild gait impairment, whereas it was 0% in both the *Mild* and *Severe* groups. Notably, the absence of awareness in the *Mild* group—a stage at which gait impairment is thought to begin to manifest—suggests that information about SM-GT may not be systematically provided even at stages when gait difficulties first become apparent. With respect to FoG severity, awareness remained around 50% in several groups (Normal: 54.5%; Slight: 50.0%; Moderate: 54.5%), whereas it was markedly higher in the *Mild* group (85.7%) and absent in the *Severe* group (0%). It should be noted, however, that sample sizes were small in some Normal and severe subcategories across all indicators, and these findings should therefore be interpreted with caution. Interventions using visual and auditory cues are recommended in clinical guidelines and reviews as important rehabilitation strategies for gait disturbances, including FoG (Domingos et al., 2018; Giorgi et al., 2024; Osborne et al., 2022; Rutz & Benninger, 2020), and are typically implemented among individuals with mild to moderate PD who are capable of independent walking. Accordingly, the particularly high awareness observed in the *Mild* FoG group may reflect the fact that individuals who are aware of emerging FoG symptoms while still maintaining functional walking are more likely to access information about SM-GT through interactions with physical therapists or other rehabilitation professionals. This pattern underscores the importance of systematically introducing SM-GT at stages when gait disturbances and FoG begin to emerge. Conversely, for individuals with severe FoG, reduced activity levels and limited opportunities to engage in rehabilitation may substantially restrict exposure to information about SM-GT (Ghielen et al., 2020; Tan et al., 2011; Zaman et al., 2021).

### Limited Implementation of SM-GT in Inpatient and Home-Based Rehabilitation

Analysis of SM-GT implementation in inpatient and home-based rehabilitation revealed limited usage, with rates of 26.1% in inpatient settings and 9.1% in home-based rehabilitation. When usage was further examined in relation to ADL level, gait and balance status, and FoG severity, implementation remained limited even among participants with moderate levels of disability—a group for whom MbM and RAS have been shown to be effective (de Dreu et al., 2012; Forte et al., 2021; Ye et al., 2022; Zhou et al., 2021). This pattern suggests that although gait rehabilitation itself is conducted on a regular basis, SM-GT has not been widely incorporated into routine rehabilitation programs, pointing to a potential gap between the accumulating evidence base and real-world clinical implementation. Survey studies involving people with PD have shown that music is frequently used in various aspects of daily life, primarily for purposes such as emotional regulation and relaxation; however, the use of music specifically to support gait has been reported to be relatively uncommon (Rose et al., 2023, 2025). Previous work has also highlighted the mechanistic link between rhythm perception and motor facilitation, supporting the potential for music to improve motor symptoms (Nombela et al., 2013). At the same time, it has been noted that RAS protocols—including stimulus parameters and training conditions—remain heterogeneous across studies, and that evidence supporting their application in home-based rehabilitation is still limited (Giorgi et al., 2024; Muthukrishnan et al., 2019).

Taken together, these findings suggest that although people with PD may have substantial potential to utilize music as a resource for gait support, the low implementation rates observed in the present study may reflect a situation in which this potential is not being fully realized in current rehabilitation practice.

### Daily-Life Use of SM-GT and Functional Status Differences

Experience of using SM-GT in daily-life contexts was reported by only 31.1% of participants, indicating that many individuals do not sufficiently utilize sound- or music-based interventions during everyday walking. This limited usage contrasts with evidence from meta-analyses and reviews demonstrating that RAS and MbM can improve gait speed, balance, and ADL (de Dreu et al., 2012; Forte et al., 2021; Zhou et al., 2021), as well as stride length (de Dreu et al., 2012; Forte et al., 2021) and suggests that implementation at the level of daily life remains inadequate. Previous studies have similarly noted that benefits observed in clinical or structured training settings do not necessarily generalize to everyday-life contexts (Lim et al., 2005; Rocha et al., 2014), and the present findings are consistent with these observations.

When daily-life use of SM-GT was examined by ADL level and gait status, higher usage rates were observed among groups with mild impairment (e.g., ADL *Mild disability*: 37.9%; gait *Mild*: 60.0%), whereas usage tended to remain at approximately 20–30% or lower among groups with very mild symptoms or with moderate to severe impairment. Interpretation of categories with extremely small sample sizes—such as the ADL *Asymptomatic* and *Moderately severe disability* groups (both n = 2) and the gait *Severe* group (n = 2)—requires caution. Overall, these distributions are broadly consistent with prior findings indicating that factors such as perceived need for exercise, physical capacity, fear of falling, and the perceived burden associated with home-based exercise influence engagement in physical activity (Ellis et al., 2013). Specifically, individuals with mild impairment may simultaneously retain awareness of gait difficulties and independent walking ability, resulting in greater exposure to rehabilitation services and non-pharmacological interventions and making SM-GT more feasible to incorporate into daily life. In contrast, individuals with very mild symptoms may not perceive a clear need for gait training, while those with moderate or greater impairment may encounter physical, psychological, and environmental barriers that make initiating or sustaining SM-GT in daily-life contexts more difficult.

Comparisons based on FoG severity revealed that daily-life use of SM-GT was highest in the *Moderate* group (45.5%, n = 11), whereas no use was reported in the *Severe* group (0%, n = 1). Although the sample size of the *Severe* group was extremely small and thus limits generalizability, previous research has shown that increasing FoG severity is associated not only with reduced gait speed and stride length but also with greater restrictions in ADL and decreased overall activity levels (Rider et al., 2025; Tan et al., 2011). Individuals with more severe FoG are therefore likely to have fewer opportunities for everyday walking and gait-related training.

Moreover, FoG has been shown to be longitudinally associated with fear of falling and anxiety (Ghielen et al., 2020), and individuals with FoG are more prone to activity avoidance as a consequence of elevated fall risk (Nonnekes et al., 2015). As a result, individuals with severe FoG may experience substantial restrictions in daily walking itself, reducing opportunities to initiate or maintain SM-GT in everyday contexts. In addition, systematic reviews of exercise, training, and cueing interventions for FoG have noted that most intervention studies target individuals with mild to moderate FoG who are capable of independent walking (Gilat et al., 2021; Kwok et al., 2022). This focus of clinical research and rehabilitation practice on patients prior to severe FoG may have contributed to relatively limited opportunities for the introduction or recommendation of SM-GT in daily life among individuals with severe FoG, consistent with the near absence of reported use in this group. Similarly, studies of gait rehabilitation using music or rhythmic auditory cues have predominantly focused on people with PD who retain independent walking ability (Murgia et al., 2018), a pattern that is consistent with the present findings. Taken together, these results suggest that, for the dissemination of SM-GT in daily-life contexts, it is realistic to primarily target individuals who retain independent walking ability and exhibit mild impairment or moderate FoG. At the same time, for individuals with very mild symptoms or with severe impairment, more individualized introduction strategies are likely to be required, including efforts to enhance awareness of the need for gait support and careful consideration of safety.

### Discrepancy Between Recognized and Utilized Auditory Cues in SM-GT

In the context of SM-GT, the metronome was the most frequently recognized auditory cue, whereas in terms of actual use, hand clapping/verbal cueing and music were more commonly employed across inpatient rehabilitation, home-based rehabilitation, and daily-life contexts, with the metronome ranking relatively lower. This discrepancy between “recognition” and “use” may reflect the longstanding positioning of the metronome as a prototypical auditory cue in conventional RAS research and clinical practice. In many intervention studies, metronomes have been widely used, and evidence—including findings from meta-analyses—has repeatedly demonstrated that RAS incorporating metronome cues is effective in improving gait speed, stride length, and balance (Ashoori et al., 2015; Bella et al., 2017; Thaut et al., 1996). Consequently, the metronome may be readily recalled at the level of conceptual recognition as a typical example of SM-GT, whereas the cues actually used in clinical and everyday settings appear to be more diverse, as suggested by the present data. Notably, the high frequency of reported use of hand clapping and verbal cueing supports the importance of selecting and adapting cues to individual patients. This finding is consistent with reports indicating that human-generated rhythmic stimuli, such as clapping or verbal rhythmic cues, are widely used in practice (Mancini et al., 2018), as well as with evidence demonstrating the utility of rhythmic clapping as an assistive strategy for gait impairment (Suteerawattananon et al., 2004). Hand clapping and verbal cueing offer several practical advantages: they require no equipment, allow flexible real-time adjustment of timing in accordance with walking speed or stride length, and can be readily combined with verbal instruction or encouragement. At the same time, their sustained use in daily-life contexts often depends on the involvement of others, such as family members or caregivers. Accordingly, an important challenge for future implementation research is to determine how cues that can be used independently—such as metronomes, portable devices, or music playlists tailored to individual preferences—can be effectively designed and disseminated without reliance on external assistance.

### Limitations

This study has several limitations related to sample characteristics. First, the distribution of participants’ ADL levels, gait ability, and FoG severity was not uniform, with a predominance of individuals with mild to moderate impairment and very few participants with severe disability. By primarily targeting individuals with mild to moderate PD—who are considered more likely to have access to SM-GT—the present study was able to provide a detailed characterization of awareness and use of SM-GT within this subgroup, which represents a key strength of the study. At the same time, the levels of awareness and use observed here largely reflect the characteristics of a relatively independent population, and caution is therefore warranted when generalizing these findings to the broader PD population, particularly to individuals with more severe motor impairment or greater ADL limitations.

Second, the present study was based on onsite survey data collected in a limited number of regions within Japan, specifically Akashi, Kobe, and Yonago. Individuals who are able to attend patient meetings or rehabilitation-related events held in these areas are likely to represent a relatively active subgroup, characterized by preserved mobility and a proactive approach to accessing information through face-to-face interactions. Accordingly, the present findings should be interpreted not as representative of the “average” PD population in Japan, but rather as reflecting patterns that may be characteristic of individuals who are relatively well positioned in terms of information access. Importantly, however, the observation that awareness and implementation of SM-GT were still limited even within this comparatively advantaged group provides a critical indication that substantial gaps remain in the dissemination of sound- and music-based interventions.

Finally, the scope of the present study was limited to Japan. One of the strengths of this study lies in its provision of the first systematic overview of the current status of sound-based interventions, including SM-GT, among people with PD in Japan, with the aim of promoting wider adoption of promising gait interventions. Future research would benefit from combining onsite and online survey methods to achieve broader generalizability through nationwide surveys that include individuals with more diverse levels of disease severity and living circumstances. Moreover, when considering international generalizability, differences in healthcare systems, rehabilitation delivery structures, and musical cultures preclude direct extrapolation of the present findings to PD populations in other countries. Accordingly, comparable investigations conducted across different national and regional contexts will be necessary.

## Conclusion

This study provides the first systematic description of awareness and use of sound- and music-based gait training (SM-GT) among people with PD in Japan. Although more than half of participants reported being aware of SM-GT, its actual implementation in inpatient rehabilitation, home-based rehabilitation, and daily-life contexts remained limited, revealing a substantial gap between awareness and use. This discrepancy was evident even among individuals with mild to moderate functional impairment, a group for whom auditory cue–based interventions have been shown to be effective.

The present findings further highlight heterogeneity in the types of auditory cues associated with SM-GT. While the metronome was most commonly recognized as a prototypical cue, hand clapping, verbal cueing, and music were more frequently used in practice, underscoring the importance of flexibility and individual adaptation in real-world gait rehabilitation. Taken together, these results suggest that SM-GT is not yet fully integrated into routine rehabilitation or daily walking support, despite a growing evidence base supporting its efficacy.

By documenting real-world patterns of awareness, use, and cue selection, this study complements existing clinical trials and offers an implementation-oriented perspective on SM-GT. These insights may inform future strategies to promote the dissemination and practical adoption of sound- and music-based interventions in both clinical and everyday settings, particularly within the Japanese context.

## Acknowledgements

We would like to sincerely thank all participants for their valuable cooperation and contributions to this study.

## Author contributions

**Conceptualization:** TS, YU, HY, SI, YM, TI, MS, SF

**Data curation:** TS, HY

**Formal analysis:** TS

**Funding acquisition:** MS, SF

**Investigation:** TS, YU, HY, SI, YM, TI, MS

**Methodology:** TS, YU, HY, SI, YM, TI, MS, SF

**Project administration:** MS, SF

**Resources:** TS, YU, HY, SI, YM, TI, MS, SF

**Software:** TS

**Supervision:** MS, SF

**Visualization:** TS

**Writing - original draft:** TS

**Writing - review & editing:** YU, HY, SI, YM, TI, MS, SF

## Statements and declarations

### Ethics considerations

This study was approved by the Ethics Committee of Reiwa Health Sciences University (approval #24-003) on 26 June 2025. This research was conducted ethically in accordance with the World Medical Association Declaration of Helsinki.

### Consent to participate

All participants received a detailed explanation of the study and signed written informed consent.

### Consent for publication

Not applicable.

### Data availability

The questionnaires, raw data, and analysis scripts used in this study are publicly available in an Open Science Framework (OSF) repository at https://osf.io/5v2fu/.

## References

Ashoori, A., Eagleman, D. M., & Jankovic, J. (2015). Effects of auditory rhythm and music on gait disturbances in Parkinson’s disease. Frontiers in Neurology, 6, 234. 10.3389/fneur.2015.00234

Barnish, M. S., & Barran, S. M. (2020). A systematic review of active group-based dance, singing, music therapy and theatrical interventions for quality of life, functional communication, speech, motor function and cognitive status in people with Parkinson’s disease. BMC Neurology, 20(1), 371. 10.1186/s12883-020-01938-3

Bella, S. D., Benoit, C.-E., Farrugia, N., Keller, P. E., Obrig, H., Mainka, S., & Kotz, S. A. (2017). Gait improvement via rhythmic stimulation in Parkinson’s disease is linked to rhythmic skills. Scientific Reports, 7(1), 42005. 10.1038/srep42005

Bella, S. D., Benoit, C.-E., Farrugia, N., Schwartze, M., & Kotz, S. A. (2015). Effects of musically cued gait training in Parkinson’s disease: beyond a motor benefit. Annals of the New York Academy of Sciences, 1337(1), 77–85. 10.1111/nyas.12651

Ben-Shlomo, Y., Darweesh, S., Llibre-Guerra, J., Marras, C., San Luciano, M., & Tanner, C. (2024). The epidemiology of Parkinson’s disease. Lancet, 403(10423), 283–292. 10.1016/S0140-6736(23)01419-8

Bock, M. A., Brown, E. G., Zhang, L., & Tanner, C. (2022). Association of motor and nonmotor symptoms with health-related quality of life in a large online cohort of people with Parkinson disease. Neurology, 98(22), e2194–e2203. 10.1212/wnl.0000000000200113

Bourdon, A., Damm, L., Dotov, D., Ihalainen, P., Dalla Bella, S., Bardy, B. G., & Cochen De Cock, V. (2025). Gait ecological assessment in persons with Parkinson’s disease engaged in a synchronized musical rehabilitation program. NPJ Parkinson’s Disease, 11(1), 12. 10.1038/s41531-024-00852-6

Bradley, M., O’Loughlin, S., Donlon, E., Gallagher, A., O’Keeffe, C., Inocentes, J., Ruggieri, F., Reilly, R. B., Walsh, R., Lynch, T., Di Luca, D. G., & Fearon, C. (2025). Determining falls risk in people with Parkinson’s disease using wearable sensors: A systematic review. Sensors (Basel, Switzerland), 25(13), 4071. 10.3390/s25134071

Calabrò, R. S., Naro, A., Filoni, S., Pullia, M., Billeri, L., Tomasello, P., Portaro, S., Di Lorenzo, G., Tomaino, C., & Bramanti, P. (2019). Walking to your right music: a randomized controlled trial on the novel use of treadmill plus music in Parkinson’s disease. Journal of Neuroengineering and Rehabilitation, 16(1), 68. 10.1186/s12984-019-0533-9

Carapellotti, A. M., Stevenson, R., & Doumas, M. (2020). The efficacy of dance for improving motor impairments, non-motor symptoms, and quality of life in Parkinson’s disease: A systematic review and meta-analysis. PloS One, 15(8), e0236820. 10.1371/journal.pone.0236820

Cochen De Cock, V., Dotov, D., Damm, L., Lacombe, S., Ihalainen, P., Picot, M. C., Galtier, F., Lebrun, C., Giordano, A., Driss, V., Geny, C., Garzo, A., Hernandez, E., Van Dyck, E., Leman, M., Villing, R., Bardy, B. G., & Dalla Bella, S. (2021). BeatWalk: Personalized music-based gait rehabilitation in Parkinson’s disease. Frontiers in Psychology, 12, 655121. 10.3389/fpsyg.2021.655121

Creaby, M. W., & Cole, M. H. (2018). Gait characteristics and falls in Parkinson’s disease: A systematic review and meta-analysis. Parkinsonism & Related Disorders, 57, 1–8. 10.1016/j.parkreldis.2018.07.008

Cronin, P., Collins, L. M., & Sullivan, A. M. (2024). Impacts of gait freeze on quality of life in Parkinson’s disease, from the perspectives of patients and their carers. Irish Journal of Medical Science, 193(4), 2041–2050. 10.1007/s11845-024-03673-x

de Dreu, M. J., van der Wilk, A. S. D., Poppe, E., Kwakkel, G., & van Wegen, E. E. H. (2012). Rehabilitation, exercise therapy and music in patients with Parkinson’s disease: a meta-analysis of the effects of music-based movement therapy on walking ability, balance and quality of life. Parkinsonism & Related Disorders, 18 *Suppl 1*, S114–S119. 10.1016/S1353-8020(11)70036-0

Delgado-Alvarado, M., Marano, M., Santurtún, A., Urtiaga-Gallano, A., Tordesillas-Gutierrez, D., & Infante, J. (2020). Nonpharmacological, nonsurgical treatments for freezing of gait in Parkinson’s disease: A systematic review. Movement Disorders: Official Journal of the Movement Disorder Society, 35(2), 204–214. 10.1002/mds.27913

Domingos, J., Keus, S. H. J., Dean, J., de Vries, N. M., Ferreira, J. J., & Bloem, B. R. (2018). The European Physiotherapy Guideline for Parkinson’s disease: Implications for neurologists. Journal of Parkinson’s Disease, 8(4), 499–502. 10.3233/JPD-181383

Dorsey, E. R., & Bloem, B. R. (2018). The Parkinson pandemic-A call to action. JAMA Neurology, 75(1), 9–10. 10.1001/jamaneurol.2017.3299

Ellis, T., Boudreau, J. K., DeAngelis, T. R., Brown, L. E., Cavanaugh, J. T., Earhart, G. M., Ford, M. P., Foreman, K. B., & Dibble, L. E. (2013). Barriers to exercise in people with Parkinson disease. Physical Therapy, 93(5), 628–636. 10.2522/ptj.20120279

Forte, R., Tocci, N., & De Vito, G. (2021). The impact of exercise intervention with rhythmic auditory stimulation to improve gait and mobility in Parkinson disease: An umbrella review. Brain Sciences, 11(6), 685. 10.3390/brainsci11060685

Gazewood, J. D., Richards, D. R., & Clebak, K. (2013). Parkinson disease: an update. American Family Physician, 87(4), 267–273. https://pubmed.ncbi.nlm.nih.gov/23418798/

GBD 2016 Parkinson’s Disease Collaborators. (2018). Global, regional, and national burden of Parkinson’s disease, 1990-2016: a systematic analysis for the Global Burden of Disease Study 2016. The Lancet Neurology, 17(11), 939–953. 10.1016/S1474-4422(18)30295-3

Ge, H.-L., Chen, X.-Y., Lin, Y.-X., Ge, T.-J., Yu, L.-H., Lin, Z.-Y., Wu, X.-Y., Kang, D.-Z., & Ding, C.-Y. (2020). The prevalence of freezing of gait in Parkinson’s disease and in patients with different disease durations and severities. Chinese Neurosurgical Journal, 6, 17. 10.1186/s41016-020-00197-y

Ghai, S., Ghai, I., Schmitz, G., & Effenberg, A. O. (2018). Effect of rhythmic auditory cueing on parkinsonian gait: A systematic review and meta-analysis. Scientific Reports, 8(1), 506. 10.1038/s41598-017-16232-5

Ghielen, I., Koene, P., Twisk, J. W., Kwakkel, G., van den Heuvel, O. A., & van Wegen, E. E. (2020). The association between freezing of gait, fear of falling and anxiety in Parkinson’s disease: a longitudinal analysis. Neurodegenerative Disease Management, 10(3), 159–168. 10.2217/nmt-2019-0028

Gilat, M., Ginis, P., Zoetewei, D., De Vleeschhauwer, J., Hulzinga, F., D’Cruz, N., & Nieuwboer, A. (2021). A systematic review on exercise and training-based interventions for freezing of gait in Parkinson’s disease. NPJ Parkinson’s Disease, 7(1), 81. 10.1038/s41531-021-00224-4

Ginis, P., Heremans, E., Ferrari, A., Dockx, K., Canning, C. G., & Nieuwboer, A. (2017). Prolonged walking with a wearable system providing intelligent auditory input in people with Parkinson’s disease. Frontiers in Neurology, 8, 128. 10.3389/fneur.2017.00128

Giorgi, F., Donati, D., & Tedeschi, R. (2024). Cueing interventions for gait and balance in Parkinson’s disease: A scoping review of current evidence. *Applied Sciences (Basel*, Switzerland), 14(24), 11781. 10.3390/app142411781

Goetz, C. G., Tilley, B. C., Shaftman, S. R., Stebbins, G. T., Fahn, S., Martinez-Martin, P., Poewe, W., Sampaio, C., Stern, M. B., Dodel, R., Dubois, B., Holloway, R., Jankovic, J., Kulisevsky, J., Lang, A. E., Lees, A., Leurgans, S., LeWitt, P. A., Nyenhuis, D., … Movement Disorder Society UPDRS Revision Task Force. (2008). Movement Disorder Society-sponsored revision of the Unified Parkinson’s Disease Rating Scale (MDS-UPDRS): scale presentation and clinimetric testing results. Movement Disorders: Official Journal of the Movement Disorder Society, 23(15), 2129–2170. 10.1002/mds.22340

Hely, M. A., Reid, W. G. J., Adena, M. A., Halliday, G. M., & Morris, J. G. L. (2008). The Sydney multicenter study of Parkinson’s disease: the inevitability of dementia at 20 years. Movement Disorders: Official Journal of the Movement Disorder Society, 23(6), 837–844. 10.1002/mds.21956

Herman, T., Barer, Y., Bitan, M., Sobol, S., Giladi, N., & Hausdorff, J. M. (2023). A meta-analysis identifies factors predicting the future development of freezing of gait in Parkinson’s disease. NPJ Parkinson’s Disease, 9(1), 158. 10.1038/s41531-023-00600-2

Kwok, J. Y. Y., Smith, R., Chan, L. M. L., Lam, L. C. C., Fong, D. Y. T., Choi, E. P. H., Lok, K. Y. W., Lee, J. J., Auyeung, M., & Bloem, B. R. (2022). Managing freezing of gait in Parkinson’s disease: a systematic review and network meta-analysis. Journal of Neurology, 269(6), 3310–3324. 10.1007/s00415-022-11031-z

Lee, H., & Ko, B. (2023). Effects of music-based interventions on motor and non-motor symptoms in patients with Parkinson’s disease: A systematic review and meta-analysis. International Journal of Environmental Research and Public Health, 20(2), 1046. 10.3390/ijerph20021046

Lim, I., van Wegen, E., de Goede, C., Deutekom, M., Nieuwboer, A., Willems, A., Jones, D., Rochester, L., & Kwakkel, G. (2005). Effects of external rhythmical cueing on gait in patients with Parkinson’s disease: a systematic review. Clinical Rehabilitation, 19(7), 695–713. 10.1191/0269215505cr906oa

Li, X., Chen, C., Pan, T., Zhou, X., Sun, X., Zhang, Z., Wu, D., & Chen, X. (2024). Trends and hotspots in non-motor symptoms of Parkinson’s disease: a 10-year bibliometric analysis. Frontiers in Aging Neuroscience, 16, 1335550. 10.3389/fnagi.2024.1335550

Mancini, M., Smulders, K., Harker, G., Stuart, S., & Nutt, J. G. (2018). Assessment of the ability of open-and closed-loop cueing to improve turning and freezing in people with Parkinson’s disease. Scientific Reports, 8(1), 12773. 10.1038/s41598-018-31156-4

Mantri, S., Ghilardi, M. F., Lessard, N., Norcini, M., Di Rocco, A., Wallock, K., Lehr, J., Okun, M. S., & World Summit Steering Committee. (2025). Proceedings of the world summit on parkinson’s disease. NPJ Parkinson’s Disease, 11(1), 293. 10.1038/s41531-025-01123-8

Matsuda, N., Takamatsu, Y., Sawada, M., & Aiba, I. (2025). Performance of a two-week rehabilitation improves motor function in inpatients with progressive supranuclear palsy: A pre-post study. Brain Sciences, 15(1), 88. 10.3390/brainsci15010088

Murgia, M., Pili, R., Corona, F., Sors, F., Agostini, T. A., Bernardis, P., Casula, C., Cossu, G., Guicciardi, M., & Pau, M. (2018). The use of footstep sounds as rhythmic auditory stimulation for gait rehabilitation in Parkinson’s disease: A randomized controlled trial. Frontiers in Neurology, 9. 10.3389/fneur.2018.00348

Muthukrishnan, N., Abbas, J. J., Shill, H. A., & Krishnamurthi, N. (2019). Cueing paradigms to improve gait and posture in Parkinson’s disease: A narrative review. Sensors (Basel, Switzerland), 19(24), 5468. 10.3390/s19245468

Nemade, D., Subramanian, T., & Shivkumar, V. (2021). An update on medical and surgical treatments of Parkinson’s disease. Aging and Disease, 12(4), 1021–1035. 10.14336/AD.2020.1225

Nielsen, T. L., Kruse, N. B., Haahr, A., Hjelle, E. G., Bragstad, L. K., Palmar-Santos, A., Navarta-Sánchez, M. V., Pedraz-Marcos, A., Pires, S. B., Roberts, H. C., & Portillo, M. C. (2022). Exploring health and social services in Denmark, Norway, Spain and the United Kingdom for the development of Parkinson’s care pathways. A document analysis. Health & Social Care in the Community, 30(6), e3507–e3518. 10.1111/hsc.13970

Nieuwboer, A., Kwakkel, G., Rochester, L., Jones, D., van Wegen, E., Willems, A. M., Chavret, F., Hetherington, V., Baker, K., & Lim, I. (2007). Cueing training in the home improves gait-related mobility in Parkinson’s disease: the RESCUE trial. *Journal of Neurology*, Neurosurgery, and Psychiatry, 78(2), 134–140. 10.1136/jnnp.200X.097923

Nombela, C., Hughes, L. E., Owen, A. M., & Grahn, J. A. (2013). Into the groove: can rhythm influence Parkinson’s disease? Neuroscience and Biobehavioral Reviews, *37*(10 Pt 2), 2564–2570. 10.1016/j.neubiorev.2013.08.003

Nonnekes, J., Snijders, A. H., Nutt, J. G., Deuschl, G., Giladi, N., & Bloem, B. R. (2015). Freezing of gait: a practical approach to management. Lancet Neurology, 14(7), 768–778. 10.1016/S1474-4422(15)00041-1

Nutt, J. G., Bloem, B. R., Giladi, N., Hallett, M., Horak, F. B., & Nieuwboer, A. (2011). Freezing of gait: moving forward on a mysterious clinical phenomenon. Lancet Neurology, 10(8), 734–744. 10.1016/S1474-4422(11)70143-0

Osborne, J. A., Botkin, R., Colon-Semenza, C., DeAngelis, T. R., Gallardo, O. G., Kosakowski, H., Martello, J., Pradhan, S., Rafferty, M., Readinger, J. L., Whitt, A. L., & Ellis, T. D. (2022). Physical therapist management of Parkinson disease: A clinical practice guideline from the American Physical Therapy Association. Physical Therapy, 102(4). 10.1093/ptj/pzab302

Perez-Lloret, S., Negre-Pages, L., Damier, P., Delval, A., Derkinderen, P., Destée, A., Meissner, W. G., Schelosky, L., Tison, F., & Rascol, O. (2014). Prevalence, determinants, and effect on quality of life of freezing of gait in Parkinson disease. JAMA Neurology, 71(7), 884–890. 10.1001/jamaneurol.2014.753

Peterson, D. S., & Horak, F. B. (2016). Neural control of walking in people with parkinsonism. Physiology (Bethesda, Md.), 31(2), 95–107. 10.1152/physiol.00034.2015

Pohl, P., Wressle, E., Lundin, F., Enthoven, P., & Dizdar, N. (2020). Group-based music intervention in Parkinson’s disease - findings from a mixed-methods study. Clinical Rehabilitation, 34(4), 533–544. 10.1177/0269215520907669

Porciuncula, F., Cavanaugh, J. T., Zajac, J., Wendel, N., Baker, T., Arumukhom Revi, D., Eklund, N., Holmes, M. B., Awad, L. N., & Ellis, T. D. (2025). Amplifying walking activity in Parkinson’s disease through autonomous music-based rhythmic auditory stimulation: randomized controlled trial. NPJ Parkinson’s Disease, 11(1), 100. 10.1038/s41531-025-00952-x

Pringsheim, T., Jette, N., Frolkis, A., & Steeves, T. D. L. (2014). The prevalence of Parkinson’s disease: a systematic review and meta-analysis: PD PREVALENCE. Movement Disorders: Official Journal of the Movement Disorder Society, 29(13), 1583–1590. 10.1002/mds.25945

Reeve, A., Simcox, E., & Turnbull, D. (2014). Ageing and Parkinson’s disease: why is advancing age the biggest risk factor? Ageing Research Reviews, 14(100), 19–30. 10.1016/j.arr.2014.01.004

Rider, J. V., Manalang, K. L. C., & Longhurst, J. K. (2025). Freezing of gait is associated with daily activity limitations among individuals with Parkinson’s disease and mild cognitive impairment. Occupational Therapy in Health Care, 39(2), 361–375. 10.1080/07380577.2024.2314181

Rocha, P. A., Porfírio, G. M., Ferraz, H. B., & Trevisani, V. F. M. (2014). Effects of external cues on gait parameters of Parkinson’s disease patients: a systematic review. Clinical Neurology and Neurosurgery, 124, 127–134. 10.1016/j.clineuro.2014.06.026

Rose, D. C., Poliakoff, E., Young, W. R., & Phillips, M. (2023). Music moves me in more ways than one: An online survey investigating the everyday use of music among people with Parkinson’s. Music & Science, 6(20592043231197792). 10.1177/20592043231197792

Rose, D. C., Stadelmann, M., Jerjen, R., Köchli, S., Senn, O., Baldassarre, A., Poliakoff, E., & Phillips, M. (2025). The use of music among Swiss people with Parkinson’s: A mixed methods survey and comparison to the UK findings. Music & Science, 8(20592043251371380). 10.1177/20592043251371380

Rutz, D. G., & Benninger, D. H. (2020). Physical therapy for freezing of gait and gait impairments in Parkinson disease: A systematic review. *PM & R: The Journal of Injury*, Function, and Rehabilitation, 12(11), 1140–1156. 10.1002/pmrj.12337

Scataglini, S., Van Bocxlaer, C., Jansen, L., Van Es, L., Van Laerhoven, C., & Truijen, S. (2025). Influence of wearable rhythmic auditory stimulation on Parkinson’s disease, multiple sclerosis, and stroke: a systematic review and meta-analysis. Scientific Reports, 15(1), 21432. 10.1038/s41598-025-05952-8

Schenkman, M., Hall, D. A., Barón, A. E., Schwartz, R. S., Mettler, P., & Kohrt, W. M. (2012). Exercise for people in early- or mid-stage Parkinson disease: a 16-month randomized controlled trial. Physical Therapy, 92(11), 1395–1410. 10.2522/ptj.20110472

Smulders, K., Dale, M. L., Carlson-Kuhta, P., Nutt, J. G., & Horak, F. B. (2016). Pharmacological treatment in Parkinson’s disease: Effects on gait. Parkinsonism & Related Disorders, 31(C), 3–13. 10.1016/j.parkreldis.2016.07.006

Su, D., Cui, Y., He, C., Yin, P., Bai, R., Zhu, J., Lam, J. S. T., Zhang, J., Yan, R., Zheng, X., Wu, J., Zhao, D., Wang, A., Zhou, M., & Feng, T. (2025). Projections for prevalence of Parkinson’s disease and its driving factors in 195 countries and territories to 2050: modelling study of Global Burden of Disease Study 2021. BMJ (Clinical Research Ed.), 388, e080952. 10.1136/bmj-2024-080952

Suteerawattananon, M., Morris, G. S., Etnyre, B. R., Jankovic, J., & Protas, E. J. (2004). Effects of visual and auditory cues on gait in individuals with Parkinson’s disease. Journal of the Neurological Sciences, 219(1-2), 63–69. 10.1016/j.jns.2003.12.007

Sveinbjornsdottir, S. (2016). The clinical symptoms of Parkinson’s disease. Journal of Neurochemistry, 139 *Suppl 1*, 318–324. 10.1111/jnc.13691

Tan, D. M., McGinley, J. L., Danoudis, M. E., Iansek, R., & Morris, M. E. (2011). Freezing of gait and activity limitations in people with Parkinson’s disease. Archives of Physical Medicine and Rehabilitation, 92(7), 1159–1165. 10.1016/j.apmr.2011.02.003

Thaut, M. H., McIntosh, G. C., Rice, R. R., Miller, R. A., Rathbun, J., & Brault, J. M. (1996). Rhythmic auditory stimulation in gait training for Parkinson’s disease patients. Movement Disorders: Official Journal of the Movement Disorder Society, 11(2), 193–200. 10.1002/mds.870110213

Thaut, M. H., Rice, R. R., Braun Janzen, T., Hurt-Thaut, C. P., & McIntosh, G. C. (2019). Rhythmic auditory stimulation for reduction of falls in Parkinson’s disease: a randomized controlled study. Clinical Rehabilitation, 33(1), 34–43. 10.1177/0269215518788615

van Swieten, J. C., Koudstaal, P. J., Visser, M. C., Schouten, H. J., & van Gijn, J. (1988). Interobserver agreement for the assessment of handicap in stroke patients. Stroke; a Journal of Cerebral Circulation, 19(5), 604–607. 10.1161/01.str.19.5.604

Wang, L., Peng, J.-L., Ou-Yang, J.-B., Gan, L., Zeng, S., Wang, H.-Y., Zuo, G.-C., & Qiu, L. (2022). Effects of rhythmic auditory stimulation on gait and motor function in Parkinson’s disease: A systematic review and meta-analysis of clinical randomized controlled studies. Frontiers in Neurology, 13, 818559. 10.3389/fneur.2022.818559

Ye, X., Li, L., He, R., Jia, Y., & Poon, W. (2022). Rhythmic auditory stimulation promotes gait recovery in Parkinson’s patients: A systematic review and meta-analysis. Frontiers in Neurology, 13, 940419. 10.3389/fneur.2022.940419

Zaman, M. S., Ghahari, S., & McColl, M. A. (2021). Barriers to accessing healthcare services for people with Parkinson’s disease: A scoping review. Journal of Parkinson’s Disease, 11(4), 1537–1553. 10.3233/JPD-212735

Zhou, Z., Zhou, R., Wei, W., Luan, R., & Li, K. (2021). Effects of music-based movement therapy on motor function, balance, gait, mental health, and quality of life for patients with Parkinson’s disease: A systematic review and meta-analysis. Clinical Rehabilitation, 35(7), 937–951. 10.1177/0269215521990526

