## Supporting Information for "Awareness and Use of Sound- and Music-Based Gait Training in People with Parkinson’s Disease in Japan: A Cross-Sectional Survey"

### A Survey Questionnaire on Gait Training Using Sound and Music for People with Parkinson's Disease

This questionnaire targets patients who have Parkinson's disease and aims to survey the awareness and frequency of use of "Sound- and music-based gait training".

Please read the content of the questions carefully before answering. If you have any questions, please feel free to ask the staff present at the venue.

Name (Signature field of the individual). ____________________

Name (Signature field for the proxy / surrogate) ____________

＊Please have the proxy / surrogate sign only if the individual finds signing difficult.

Date of birth ______________ （Age: ________） Sex: M ・ F

Prefecture of Residence ______________________________

Please fill in information regarding Parkinson's disease.

Onset: ____________________ Diagnosis: ________________

Other illness ______________________________________

PD Medication. Name: ____________ Frequency: ( ) times per day

Name: ____________ Frequency: ( ) times per day

Name: ____________ Frequency: ( ) times per day

Name: ____________ Frequency: ( ) times per day

Name: ____________ Frequency: ( ) times per day

We will ask about your condition in daily life. Please circle the condition that best describes your usual state (when medication is not effective).

1. **Asymptomatic**: No symptoms at all
2. **Symptomatic without disability:** No significant disability despite symptoms; able to carry out all usual duties and activities
3. **Slight disability**: unable to carry out all previous activities, but able to look after own affairs without assistance
4. **Moderate disability**: requiring some help, but able to walk without assistance
5. **Moderately severe disability**: unable to walk and attend to bodily needs without assistance

We will ask about your walking or balance condition. Please circle the condition that best describes your usual state (when medication is not effective).

Over the past week, have you usually had problems with balance and walking?

0: **Normal**: Not at all (no problems).

1: **Slight**: I am slightly slow or may drag a leg. I never use a walking aid.

2: **Mild**: I occasionally use a walking aid, but I do not need any help from another person.

3: **Moderate**: I usually use a walking aid (cane, walker) to walk safely without falling. However, I do not usually need the support of another person.

4: **Severe**: I usually use the support of another person to walk safely without falling.

We will ask about Freezing of Gait (FoG). Please circle the condition that best describes your usual state (when medication is not effective).

Over the past week, on your usual day when walking, do you suddenly stop or freeze as if your feet are stuck to the floor?

0: **Normal**: Not at all (no problems)

1: **Slight**: I briefly freeze, but I can easily start walking again. I do not need help from someone else or a walking aid (cane or walker) because of freezing.

2: **Mild**: I freeze and have trouble starting to walk again, but I do not need someone's help or a walking aid (cane or walker) because of freezing.

3: **Moderate**: When I freeze I have a lot of trouble starting to walk again and, because of freezing, I sometimes need to use a walking aid or need someone else's help.

4: **Severe**: Because of freezing, most or all of the time, I need to use a walking aid or someone's help.

### Sound- and Music-based Gait Training (SM-GT)

It is known that playing a consistent rhythmic sound (such as a metronome or hand clapping) while walking can make walking easier. Such sound prompts are called "auditory cues" and can lead to improved walking rhythm and stability.

Furthermore, walking in rhythm with music helps regulate the walking tempo. Using preferred music makes the activity enjoyable and is said to lead to improvements in walking rhythm and stability. In this questionnaire, we refer to this content as **"Sound- and Music-based Gait Training"**.

From here, we will ask about **"Sound- and Music-based Gait Training"**. Unlike music therapy which involves singing or dancing to music, we will only ask about walking training where sound or music is used as a cue for walking.

Please ask if you have any unclear points.

Q1. Are you aware of Sound- and Music-based Gait Training? Please circle the applicable response. If you are aware, please select / write the sound or music used.

1. Yes, I am aware **(Please select all items used and proceed to Q2.)**

☐ Metronome ◻ Hand clapping / Verbal cueing

☐ Music (Walking in rhythm with music) ◻ Other：_____________________

2. No, I am not aware **(Please proceed to Q3.)**

Q2. **(For those who selected "1: Yes, I am aware" in Q1 only)**

What was the setting where you learned about Sound- and Music-based Gait Training? Please circle the most applicable response.

1: Rehabilitation setting at the hospital

2: Examination setting by a doctor at the hospital

3: Rehabilitation setting at home

4: Books

5: Website

6: Other (Please describe below)

______________________________________________

Next, we will ask about the **"Inpatient Rehabilitation Setting"**.

You will answer about the home rehabilitation setting and daily life setting later, so please limit your answers here to the inpatient rehabilitation setting.

Q3. How often did you perform walking training during your hospitalization?

The use of sound is not relevant here. Please circle the frequency of walking training that is most applicable.

1. Daily
2. 2-3 times a week
3. Once a week
4. Once every two weeks
5. Less frequently than that

Q4. Have you had experience performing Sound- and Music-based Gait Training during the walking training you received while hospitalized? Please circle the applicable response. If you have experience, please select/write the sound or music used.

1. Yes I have **(Please select all items used and proceed to Q5.)**

   ☐ Metronome　　□ Hand clapping / Verbal cueing

   ☐ Music (Walking in rhythm with music)　□ Other____________________

2. No, I have not **(Please proceed to Q7.)**

**Q5. <u>(For those who selected "1: Yes, I have" in Q4 only)</u>**

How often did you perform Sound- and Music-based Gait Training during your hospitalization? Please circle the most applicable response.

1. Daily
2. 2-3 times a week
3. Once a week
4. Once every two weeks
5. Less frequently than that

Q6. **<u>(For those who selected "1: Yes, I have" in Q4 only)</u>**

Compared to walking training without the use of sound or music, were you able to feel a greater effect when performing Sound- and Music-based Gait Training? Please circle the most applicable response.

1. I felt a very strong effect
2. I felt an effect
3. I felt a slight effect
4. I did not feel much of an effect
5. I did not feel an effect
6. I felt absolutely no effect

Next, we will ask about the **"Home Rehabilitation Setting implemented with professionals such as physical therapists and occupational therapists"**.

We will ask about the daily life setting later, so please limit your answers here to the home rehabilitation setting.

Q7. In home rehabilitation implemented with professionals such as physical therapists and occupational therapists, how often did you perform walking training?

The use of sound is not relevant here. Please circle the frequency of walking training that is closest to your experience.

1. Daily
2. 2-3 times a week
3. Once a week
4. Once every two weeks
5. Less frequently than that

Q8. Have you had experience performing Sound- and Music-based Gait Training during home rehabilitation implemented with professionals such as physical therapists and occupational therapists?

Please circle the applicable response. If you have experience, please select / write the sound used.

1. Yes I have **(Please select all items used and proceed to Q9.)**

☐ Metronome ☐ Hand clapping / Verbal cueing

☐ Music (Walking in rhythm with music) ☐ Other____________________

2. No, I have not **(Please proceed to Q11.)**

Q9. **(For those who selected "1: Yes, I have" in Q8 only)** How often did you perform Sound- and Music-based Gait Training during home rehabilitation implemented with professionals such as physical therapists and occupational therapists? Please circle the most applicable response.

1. Daily
2. 2-3 times a week
3. Once a week
4. Once every two weeks
5. Less frequently than that

Q10. **(For those who selected "1: Yes, I have" in Q8 only)**

Compared to walking training without the use of sound or music, were you able to feel a greater effect when performing Sound- and Music-based Gait Training?

Please circle the most applicable response.

1. I felt a very strong effect
2. I felt an effect
3. I felt a slight effect
4. I did not feel much of an effect
5. I did not feel an effect
6. I felt absolutely no effect

Next, we will ask about the "Daily Life Setting". Please answer regarding content limited to your usual daily life, excluding rehabilitation.

**Q11.** This question is intended to check whether you have read and understood the questions properly. **Do not circle any option for this question.**

1. ①

2. ②

3. ③

Q12. Do you use sound or music to assist your walking in your daily life? Please circle the applicable response. If you have experience, please select / write the sound used.

1. Yes I have **(Please select all items used and proceed to Q13.)**

    ☐ Metronome ◻ Hand clapping / Verbal cueing

    ☐ Music (Walking in rhythm with music) ◻ Other______________________

2. No, I have not **(Please proceed to Q16.)**

Q13. **<u>(For those who selected "1: Yes, I have" in Q12 only)</u>**

How often do you use sound or music to assist your walking in your daily life?

Please circle the most applicable response.

1. **Use daily, without fail**: Always used regardless of symptoms
2. **Use almost daily**: Used most of the time when needed
3. **Use occasionally**: Sometimes used when needed
4. **Use sometime**s: Used rarely
5. **Use Hardly at all**: often not used even when needed

Q14. **<u>(For those who selected "1: Yes, I have" in Q12 only)</u>**

Are you currently still using sound or music for the improvement of your walking in daily life, excluding rehabilitation settings? Please circle the most applicable response.

1. Used **<u>within approximately one week of the date</u>** of this questionnaire
2. Used **<u>within approximately one month of the date</u>** of this questionnaire
3. Used **<u>within approximately six months of the date</u>** of this questionnaire
4. Used **<u>within approximately one year of the date</u>** of this questionnaire

Q15. **<u>(For those who selected "1: Yes, I have" in Q12 only)</u>**

When using sound or music to assist your walking, were you able to feel an effect? Please circle the most applicable response.

1. I felt a very strong effect
2. I felt an effect
3. I felt a slight effect
4. I did not feel much of an effect
5. I did not feel an effect
6. I felt absolutely no effect

Q16. **<u>(For those who selected "1: Yes, I have" in Q12 only)</u>**

In daily life, when using sound or music to assist your walking, who provides the sound or music cues? Please circle the most applicable response.

1. Your family member(s)
2. Yourself
3. Other person (please write below)

________________________________________

Q17. If there were shoes and an app that played sound or music synchronized with your walking to assist walking difficulty, would you want to use them now or in the future in your daily life?

1. Definitely want to use
2. Moderately want to use
3. Neither agree not disagree (Neutral)
4. Do not want to use much
5. Absolutely do not want to use

Q18. Please write freely about the use of sound or music to assist your walking in daily life, including ease of use, the sensation felt when using it, or any requests.

This concludes the questionnaire. Thank you for your answers.
